# A Methodology to Evaluate the Efficacy and Patient-Centeredness of a Patient-Facing AI Chatbot: Development and Validation of a Standardized-Patient Method

**DOI:** 10.64898/2026.10.08.26364702

**Authors:** Tauseef I. Mamun, Ananda Rou Amogh, Jennifer W. Rhodes, William Martinez, Sarah Stern, Kaustav Shah, Kevin B. Johnson, Matthew B Weinger

**Author notes:** Address correspondence to: Tauseef Mamun, PhD, Department of Biomedical Informatics Vanderbilt University Medical Center 2525 West End Avenue, Suite 1475.

## Abstract

**Background:** Patient-facing AI intake chatbots can enter clinical use without regulatory review, so their errors may first surface with actual patients.

**Objective:** To develop and pilot a remote, voice-based standardized patient (SP) method for evaluating an intake chatbot’s history-taking, documentation, and patient-centeredness.

**Methods:** Four primary care cases had Easy and Difficult variants. In Phase 1, five SPs completed 40 encounters with internal medicine residents rating portrayal fidelity. In Phase 2, SPs completed 80 encounters with a ChatGPT 4o chatbot, recorded whether critical elements were asked about and addressed satisfactorily, and rated summaries; four debriefing interviews were analyzed. In Phase 3, two blinded clinicians rated transcripts and summaries from 101 encounters (32 resident-led, 69 chatbot-led).

**Results:** Portrayal fidelity averaged 4.52 of 5. The chatbot asked about 59.3% of critical elements in Easy versus 50.2% in Difficult variants (adjusted OR 1.49; 95% CI, 1.15 to 1.93) and addressed 73.3% versus 64.6% (OR 1.47; 1.12 to 1.93); near-ceiling summary quality did not differ. SPs reported missed information and rushed pacing. Reviewer agreement was moderate to substantial for element judgments (AC1 0.49 to 0.70) but poor for totals (ICC 0.10 and 0.21). In exploratory comparisons, residents asked about more elements (adjusted difference 12.5 percentage points; 95% CI, 7.0 to 18.1) and satisfactorily addressed more (9.2 points; 3.8 to 14.6); totals did not differ.

**Conclusions:** SP portrayal was consistent enough for pre-clinical chatbot evaluation, and the method detected lower coverage in harder cases. Documentation ratings alone may miss conversational gaps, which require direct measurement.

## Introduction

Healthcare artificial intelligence (AI) is increasingly in direct contact with patients. In this paper, we describe a pre-clinical evaluation of an AI chatbot that performs clinical intake to summarize a patient’s presenting complaints for a primary care provider. Under current federal regulations, such a software tool could be marketed and used clinically without any formal federal review or evaluation. If a formal evaluation did occur, it would be on actual patients in the clinical environment. We sought to develop a pre-clinical methodology using standardized patients (SPs) to conduct a formal evaluation before clinical use.

Some patient-facing AI tools do require formal oversight. For example, algorithms embedded in medical devices and software that recommends medical treatments to patients would go through premarket Food and Drug Administration (FDA) review for safety and efficacy.^1^ But a large and growing class of patient-facing tools does not. Medical AI tools can avoid strict regulation, for example, by avoiding explicit medical claims or presenting themselves as general-wellness products. Software for medical documentation also falls outside FDA oversight. The path for oversight may be related more to how an AI tool is labeled and used than to its technical design.^1^ Moreover, there is no standardized framework for evaluating such tools after they are in clinical use. AI voice chatbots that take a patient history sit squarely in this gap, being able to be used for patient care with no formal pre-market or post-market review or evaluation.

Chatbots have been deployed to collect clinical data directly from patients,^2^ and other conversational medical systems have been deemed ready for clinical use under physician supervision.^3^ In a randomized trial of 2,069 patients across 24 medical disciplines, a patient-facing Large Language Model (LLM) conducted pre-consultation history-taking, generated preliminary diagnoses, and produced referral reports. The LLM application reduced specialist consultation time by nearly 30% while improving patient-reported communication.^4^

Existing approaches emphasize different aspects of patient-facing AI, which results in fragmented evaluation practices. A recent agentic-AI system took histories from simulated patients and was scored by blinded physicians against gold-standard case histories using a predefined rubric^5^. Its evaluation centered on the accuracy of the captured history, so portrayal fidelity, documentation quality, and the patient’s experience fell outside the study’s scope. One published framework scored a health-coach chatbot for quality and safety, where the ratings came from trained reviewers rather than the patient in the conversation^6^. A review in dermatology catalogued over 2,500 performance scores across 88 studies and found that model development had outpaced the methods meant to track it, so that evaluation designs are inconsistent and benchmarks age quickly^7^. Prior studies are limited in evaluating a voice-based patient intake chatbot on both the effectiveness of data capture and the patient experience of using it.

Accuracy is important, but it does not capture everything a good intake encounter is supposed to do. Patients judge clinical encounters heavily on whether they feel listened to and understood. Prior work treats cognitive empathy and shared understanding as genuine elements of patient-AI communication.^8^ A systematic review and meta-analysis found that chatbots often match or exceed clinicians on rated empathy,^9^ but that evidence rested almost entirely on text-only responses and scoring by outside raters rather than the patient. The authors advocated for direct patient evaluation and voice-based testing. In the present study, we used voice-based discussions and rated the quality and completeness of the interaction by the SPs who took part.

SPs offer a way to close several of these gaps. Trained SPs portray clinical cases consistently enough to support reproducible evaluation,^10,11^ which makes them a controlled and lower-cost substrate for testing an intake chatbot remotely, without a patient trial. Here, we describe and pilot test an SP-based, remote, voice-delivered method for assessing how well a patient-facing AI chatbot captures the essential elements of a chief complaint. The method rates the captured clinical content and the patient-centeredness of the encounter, and it offers a practical way to evaluate such tools before they reach patients. The study had three aims, which were addressed in three linked phases. First, we developed a consistent SP method for presenting chief complaints and examined how closely SPs followed the written scripts. Second, we tested whether the method could detect differences in chatbot performance between easier and more difficult cases. Third, we compared chatbot-led and resident-led encounters using blinded clinician review.

## Methods

### Study Design

We developed and pilot-tested a pre-clinical method to evaluate how well a patient-facing AI chatbot captures the essential elements of a patient’s chief complaint. Trained SPs interact with the chatbot remotely by voice, so the chatbot’s history-taking and documentation can be assessed without enrolling actual patients. This feasibility study was designed to establish and validate the approach rather than to judge the specific chatbot conclusively. The study was reviewed by the Vanderbilt University Medical Center Human Subject Protection Program and deemed Exempt.

The study ran in three phases (**Figure 1**). Phase 1 developed a reproducible SP method for delivering patient chief complaints with high fidelity and validated portrayal accuracy against the written scripts, with internal medicine residents serving as medical testers (MTs) in place of the chatbot. Phase 2 assessed how well the chatbot captured and documented the essential elements of each complaint across two levels of case difficulty. Phase 3 compared the chatbot-led encounters from Phase 2 with the resident-led encounters from Phase 1 through blinded review by independent internal medicine fellows.

**Figure 1.**
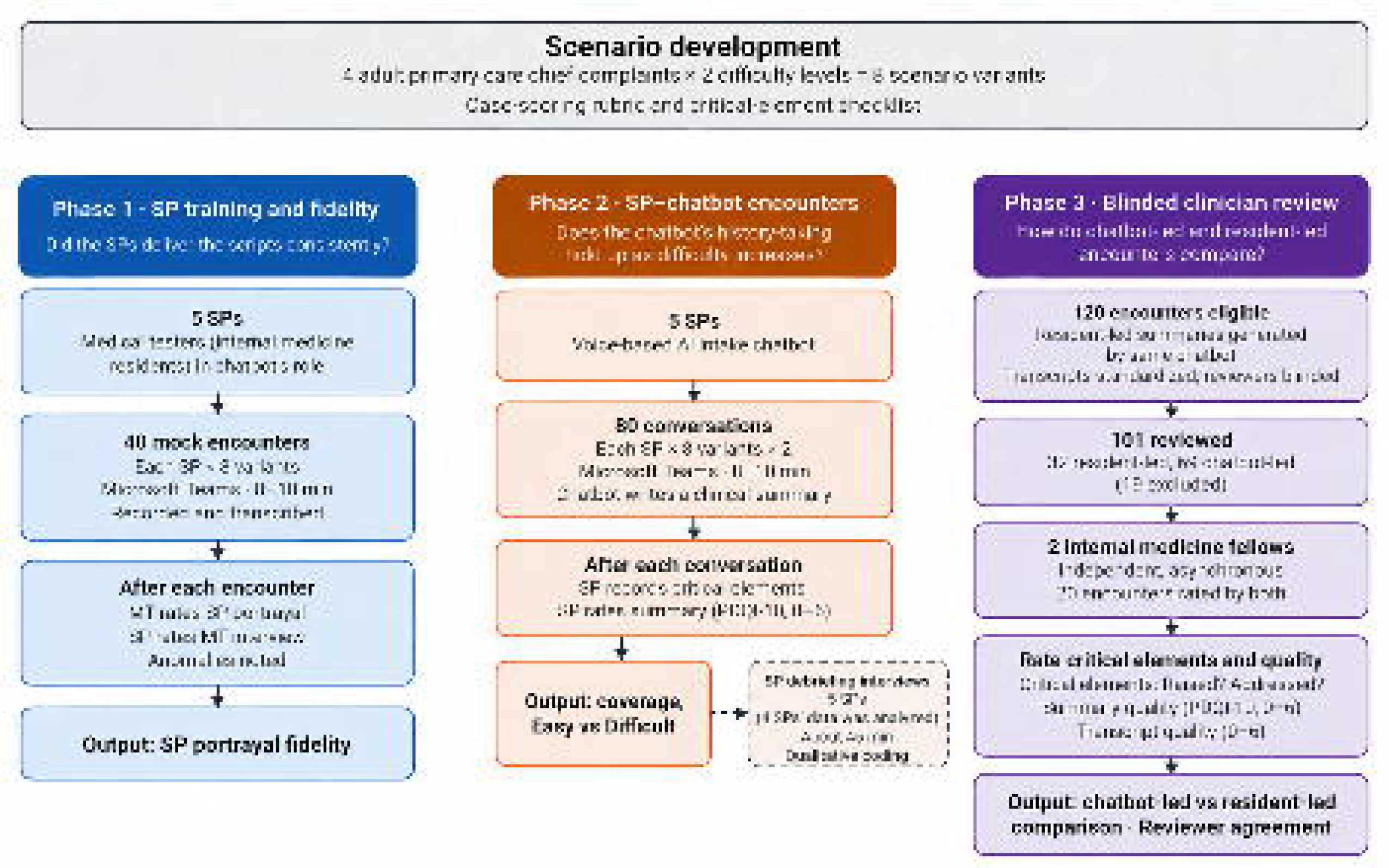
Flow of the Study

We evaluated the chatbot’s history-taking with SPs, individuals trained to portray a specific patient presentation accurately and consistently.^10,11^ Recent studies have taken a similar approach with simulated patients.^5,12–14^ In one of those studies, physicians and medical students played patients from published constructed cases, while blinded physicians rated the chatbot’s performance.^5^ Our method extends that design to voice delivery, adds explicit validation of portrayal fidelity, and rates the written summary with a documentation-quality instrument alongside the SP’s own rating of the encounter.

### The Chatbot

The AI chatbot studied, OpenAI ChatGPT 4o™ (see **Appendix A** for the chatbot’s guardrails), was a voice-based primary-care agent that acts as a virtual adult primary care intake nurse. It conducts the initial screening conversation and generates a clinical summary of the encounter for use by the primary care provider (PCP). The intent is to streamline the clinic visit and to ensure that the PCP meets the patient’s care needs.

### Scenario (Case) Development

We developed four adult primary care chief-complaint scenarios, each expected to take about 10 minutes to deliver: women’s health (abnormal uterine bleeding), mental health (anxiety disorder), an acute condition (fever and sore throat), and chronic-disease management (hypertension). The team built the cases by consensus. For this pilot, we intentionally avoided emergency conditions, overly complicated cases, and content that was highly emotional or controversial. Each scenario was initially developed as an Easy (E) variant, which presented the complaint straightforwardly, and this was then modified to create a Difficult (D) variant, which added an unfocused narrative, heightened patient emotion and/or interruptions or distraction. See **Appendix B** for Detailed Case Scripts.

To assess the case scripts, the research team built a case-scoring rubric by consensus, rating each scenario from 0 to 3 on eleven criteria covering medical realism, inclusion of the predefined critical clinical elements, patient context, the balance of relevant and distracting detail, and the cues given to the SP for emotion, pacing, and delivery (see **Appendix C** for the rubric). Two study team physicians and a non-study team physician independently scored the written case materials using the rubric. This led to a few minor refinements of each script, validated by team consensus.

The primary case author developed a checklist of case-specific clinical elements for each scenario. These were then refined by the common practice of defining critical performance elements by expert consensus^15^. Cases 1 and 2 had 12 and 13 elements, respectively, across both difficulty levels (i.e., the added difficulty of the D variant was intended to arise from the SP’s delivery rather than key clinical/management content). Case 3 had 12 elements in the E variant and 13 in D, the additional element covering financial and insurance concerns. Case 4 had 9 in the E and 16 in the D with the latter adding a positional-vertigo work-up alongside the hypertension follow-up, as well as caregiver strain and intermittently missed medication doses. This variation allowed us to test the chatbot across difficulty levels using cases that included both shared and distinct critical elements. All the surveys in the phases were done in REDCap™ (Vanderbilt University Medical Center, Nashville, TN).

#### Phase 1. SP Training and Portrayal Fidelity Validation

We trained five SPs to deliver all eight scenario variants according to widely accepted SP training practices.^16^ Each SP spent 4-5 hours (over two sessions) training with a Standardized Patient educator. Training focused on case symptoms and patient affect and portrayal as well as the differences between the Easy and Difficult variant portrayals. SPs were expected to have the same opening line (chief complaint) verbatim and to respond in the same way to elements that are important to the diagnosis. The training goal was that all five SPs be standardized and consistent. Each SP delivered all eight variants once, in a block-randomized order of four per session constrained so that variants of the same scenario never followed one another. This produced 40 encounters across ten sessions with one SP per session.

Four MTs (all internal medicine residents, referred to here as medical testers) portrayed the AI chatbot in mock encounters, again acting as an intake clinician. The MTs had a training session and reviewed the written rules that were used to train the chatbot. Three MTs conducted 8 and one MT did 16 encounters. Each encounter was conducted over Microsoft Teams, lasted 8 to 10 minutes, and was audio recorded and automatically transcribed.

Both the MT and SP rated the experience after every encounter using 5-point scales, except for the MT’s case-difficulty item, which offered four response options. Structured rating scales of this kind are an accepted way to evaluate SP portrayal accuracy^17^. The MT rated the SP’s portrayal on realism, quality of presentation, delivery style, demeanor and emotional tone, and clarity and completeness, each anchored from very low to very high. The MT also rated the difficulty of extracting and summarizing the case’s essential clinical elements (see **Appendix B**). The SP rated the MT’s interview on thoroughness, usefulness, organization, succinctness, and bias, each anchored from not at all true to very true. The SP also rated the MT’s overall quality and the difficulty of presenting the scripted case accurately and completely. Both recorded any anomalies; The MT noted whether any SP response was unreasonable or surprising, and the SP responded to three free-text prompts about disruptions, distractions, and unexpected responses.

#### Phase 2. SP-Chatbot Encounters

Each of the five SPs completed 16 conversations with the chatbot, delivering each of the eight scenario variants twice, yielding 80 conversations in total. Encounters were 8 to 10 minutes over Microsoft Teams. The chatbot produced a clinical summary within roughly two minutes after each encounter. The SP completed a structured assessment immediately after each conversation. First, the SP recorded whether the chatbot asked about each critical clinical element and whether it gave the SP enough opportunity to address that element satisfactorily. Both were rated as ‘yes’ or ‘no’, with optional free-text entry.

The SP then rated the chatbot’s clinical summary using the original PDQI-10 (0-to-6 scale with anchoring text for 0, 3, and 6 ratings) and provided narrative comments for any perceived anomaly. Each item included anchored text describing the rating criteria to reduce confusion.

The PDQI-10 rates the descriptive quality of documentation rather than whether specific content is present.^18^ It modifies the validated 9-item PDQI-9 for AI-generated documentation,^19^ eliminating the original “up-to-date” item and adding “free from hallucinations” and “free from biases.”

##### SP Debriefing Interview

All SPs from Phase 2 took part in a semi-structured debriefing interview^20^ (see **Appendix D)** of about 45 minutes after one randomly chosen session of four encounters. Data from four SPs were analyzed because transcripts were unavailable for the fifth SP. The debriefing drew on cognitive-empathy constructs (shared knowledge, shared decision-making, shared sensemaking, and common ground) that prior work links to patient-AI communication^8^ to create prompts about whether the SP and the chatbot reached mutual understanding; it also included general questions about overall satisfaction, usability, and pace.

The first author developed a 32-code codebook inductively. Two coders experienced in qualitative methods used it to independently identify and code relevant excerpts of the four debriefing transcripts. Intercoder reliability was assessed using Gwet’s AC2^21^, weighting agreement as full (1), partial (0.5), or none (0). After reliability was determined, the first coder adjudicated partial agreements and disagreements^22^.

#### Phase 3. Blinded Clinician Review of Transcripts and Summaries

Phase 3 compared chatbot-led with resident-led encounters using the transcript and the clinical summary. As the MTs did not write a clinical summary after each encounter, and to facilitate direct comparisons of the encounter content, we presented the transcript of each MT-SP encounter to the same chatbot and asked it to generate a clinical summary in its own format. A clinician standardized the opening and closing language of all SP-MT transcripts to match the chatbot transcripts. The clinician also removed filler words, pauses, and speech dysfluencies to blind the clinician reviewers to encounter type. Reviewers were also blinded to case difficulty.

Two clinicians (board-eligible internal medicine fellows) received an orientation, studied all eight case scripts and detailed written instructions, and practiced on a sample encounter with feedback. They then reviewed all available encounter transcripts and case summaries, randomly assigned, independently, and asynchronously.

Transcripts and summaries from all 120 encounters, 40 resident-led from Phase 1 and 80 chatbot-led from Phase 2, were eligible for review. Twenty encounters were assigned to both reviewers to support the reliability analysis.

For every pre-specified critical clinical element, reviewers recorded whether the encounter raised it and whether it was addressed adequately, and flagged any extraneous topics the provider introduced. Summary quality was rated on the ten PDQI-10 items using a 0-to-6 scale, summing to a total of 60. Transcript quality was rated for five items using the same scale, summing to 30: three describing the provider (realism, clarity and actionability, risk of patient harm) and two describing the SP (coverage, validity). The risk-of-harm item was rated from 0 for high risk to 6 for low risk (a higher score indicates a safer encounter.

### Statistical Analysis

We conducted all analyses in R.

The three Phases addressed different questions. Phase 1 assessed whether the SPs delivered the scripts consistently enough for their ratings to be used as a measurement instrument. Phase 2 assessed whether the chatbot’s history-taking and documentation remained consistent as case difficulty increased. Phase 3 assessed whether independent clinicians agreed when rating the same encounter. We then used those ratings to compare chatbot-led and resident-led encounters.

Encounters were grouped by Phase rather than by allocation. Resident-led encounters were conducted in Phase 1, and chatbot-led encounters were conducted in Phase 2. As the resident-led sample was smaller, comparisons between the two groups were exploratory. Effect sizes and confidence intervals were given more interpretive weight than p values.

**Phase 1.** The training phase was designed to assess whether the SPs delivered the scripts accurately. The fidelity analysis was therefore descriptive. Each portrayal item was summarized using the mean, standard deviation (SD), median, range, and proportion of ratings of 4 or higher. A per-encounter fidelity score was calculated by averaging the five portrayal items. The four SP-rated interview-quality items analyzed (thoroughness, usefulness, organization, and succinctness) were summarized similarly; the fifth, on bias, was reported separately because its label and question stem pointed in opposite directions. Each difficulty item was summarized separately by mean, SD, and range. Cronbach’s alpha was used to estimate internal consistency for the five portrayal items and the four interview-quality items. Fidelity was also examined by scenario, variant, SP, and MT.

**Phase 2.** We included records only if every checklist item was completed. Logistic regression was used to examine two outcomes for each critical element: whether the chatbot asked about it and whether the SP had enough opportunity to address it satisfactorily. Difficulty group was the main predictor, and both models adjusted for case. Standard errors were clustered by SP. Two sensitivity analyses followed. Both primary models, for elements asked and for elements addressed satisfactorily, were refitted as mixed-effects models with a random intercept for SP, and then without the case adjustment. Coverage was summarized separately for each of the four cases. Each PDQI-10 item was compared between difficulty groups using Wilcoxon rank-sum tests, with each conversation treated as one observation. Negative sentiment in the SPs’ free-text reflections was analyzed separately; each scored word was treated as one observation, standard errors were clustered by conversation, and case was not included in the model.

**Phase 3.** The data were organized so that each reviewer had one record for each encounter. We retained each reviewer’s most recent rating and calculated total scores using a complete-case rule^23^. Reviewer pairing followed the pre-specified assignment plan. Two analytic samples were used. The reliability sample included encounters rated by both reviewers and contained 260 critical-element judgments. The comparison sample included all rated encounters.

Agreement on each of the fifteen 0-to-6 items was assessed using Gwet’s AC2, a chance-corrected coefficient computed with quadratic weights^21^. We also reported exact agreement, agreement within one point, and the mean signed difference between reviewers. Agreement within one point and the mean signed difference were used to distinguish systematic rater shifts from less structured disagreement. Agreement on the two total scores was assessed using ICC(A,1), a two-way absolute-agreement single-rater intraclass correlation.

Binary critical-element judgments were assessed using Gwet’s AC1, the unweighted form of the same coefficient, and percent agreement. These measures were calculated for the pooled sample. The pooled confidence interval was estimated using a cluster bootstrap with 2,000 resamples.

Encounters rather than individual element rows were resampled. Agreement on adequacy was calculated in three ways: across all elements, across only elements that at least one reviewer marked as asked, and across the remaining elements that neither reviewer marked as asked. The second was treated as the primary and more interpretable measure. Agreement coefficients were interpreted using the Landis and Koch benchmarks^24^.

Four encounter-level outcomes were compared between the resident and chatbot groups: the PDQI-10 summary total, transcript total, proportion of critical elements asked, and proportion addressed satisfactorily. The primary analysis retained one row per reviewer per encounter. Each outcome was regressed on group, case variant, and reviewer. Reviewer was included because the two reviewers differed in leniency and were unevenly distributed across groups. We used cluster-robust CR2 standard errors at the encounter level to prevent encounters rated by both reviewers from being treated as two independent observations. Case adjustment included all eight variants while holding difficulty constant. The chatbot group was the reference level, so differences are reported as resident minus chatbot, and a positive value indicates a higher score for the resident group.

A descriptive analysis averaged the two reviewer scores within each encounter. We reported means, standard deviations, Hedges’ g, and case-adjusted contrasts. All 15 individual items were compared between groups using a single regression model fitted across items. The model included a separate intercept and group contrast for each item and adjusted for case variant and reviewer. Standard errors were cluster-robust (CR2) at the encounter level. Thus, the 30 ratings contributed by an encounter rated by both reviewers were treated as one independent unit rather than as 30 independent observations.

An omnibus test evaluated the null hypothesis that no item differed between groups. Because the items were tested jointly, this test required no correction for multiplicity. A common effect across all 15 items was also estimated. A further test assessed whether the 15 group contrasts were equal to one another, which determines whether a single common effect can adequately represent the item-level results. Two refits, with item-specific reviewer terms and with item-specific reviewer and difficulty terms, tested whether those adjustments could be treated as common across items. The 15 item-specific contrasts still represented 15 separate comparisons and were therefore corrected using the Bonferroni method^25^, which was selected for its simplicity and familiarity. An item was considered to survive correction only if it met the adjusted significance threshold. The Phase 2 comparison of Easy and Difficult cases was planned in advance as part of Aim 2. The remaining analyses were not planned in advance and are exploratory. Correction was nonetheless applied to the 15-item contrasts, to keep the item-level claim conservative rather than because the family of tests had been specified beforehand. We also compared Easy and Difficult variants in the Phase 3 clinician ratings to check that the difficulty labels worked as intended. Finally, we grouped the five transcript items into provider-rated and SP-rated subsets to check whether the two moved together.

## Results

### Phase 1. SP portrayal fidelity

Both rating instruments were completed for all 40 training encounters resulting in 40 MT ratings of SP portrayal and 40 SP ratings of MT interview quality. The five portrayal items had a mean score of 4.52 on the 5-point scale. Mean scores were 4.50 (SD 0.55) for realism, 4.40 (SD 0.71) for quality of presentation, 4.60 (SD 0.55) for delivery, 4.68 (SD 0.47) for demeanor and emotional tone, and 4.42 (SD 0.71) for clarity and completeness. Overall, 94% of portrayal ratings were 4 or higher. Every item had a median score of 5, and no MT rated any SP response as unreasonable or surprising (**Table 1**, Panel A).

**Table 1.** Phase 1 ratings using the 1-to-5 training scale (n = 40 encounters). Panel A shows MT ratings of SP portrayal. The five portrayal items had a mean score of 4.52, and 94% of portrayal ratings were 4 or higher. Panel B shows SP ratings of MT interview quality.

| Rating item | n | Mean | SD | Media<br>n | Min | Ma<br>x | ≥4 (%) |
| --- | --- | --- | --- | --- | --- | --- | --- |
| **Panel A. MT ratings of SP portrayal (fidelity)** |  |  |  |  |  |  |  |
| Realism | 40 | 4.50 | 0.55 | 5 | 3 | 5 | 97.5 |
| Quality of presentation | 40 | 4.40 | 0.71 | 5 | 3 | 5 | 87.5 |
| Delivery | 40 | 4.60 | 0.55 | 5 | 3 | 5 | 97.5 |
| Demeanor and emotional<br>tone | 40 | 4.68 | 0.47 | 5 | 4 | 5 | 100.0 |
| Clarity and completeness | 40 | 4.42 | 0.71 | 5 | 3 | 5 | 87.5 |
| **Panel B. SP ratings of MT interview quality** |  |  |  |  |  |  |  |
| Thorough | 40 | 4.55 | 0.55 | 5 | 3 | 5 | 97.5 |
| Useful | 40 | 4.42 | 0.55 | 4 | 3 | 5 | 97.5 |
| Organized | 40 | 4.70 | 0.46 | 5 | 4 | 5 | 100.0 |
| Succinct | 40 | 4.78 | 0.42 | 5 | 4 | 5 | 100.0 |

The SPs’ ratings of the MTs suggested that the mock encounters were of good fidelity. SPs rated the interviews highly on thoroughness, usefulness, organization, and succinctness, with at least 97.5% of ratings at 4 or higher on every item (Table 1, Panel B). They rated overall provider quality at 4.25. Both sets of difficulty ratings fell toward the low end of their scales. SPs rated the difficulty of presenting the case at 1.98 (SD 1.05) on a 5-point scale, and MTs rated the difficulty of extracting and summarizing it at 2.08 (SD 0.89) on a 4-point scale. The two items cover different tasks on different scales and are not directly comparable.

The SP’s “free-from-biases” item showed a different pattern with responses concentrated at both ends of the scale – 27 ratings of 5, 13 ratings of 1, and no ratings in between. This question was confusing: The rating scale made 5 a favorable score, while the question stem made 5 an unfavorable one, so raters were confused. We excluded this item from the interview-quality summary and revised both the item and the rating scale for Phases 2 and 3.

Fidelity was consistent across scenarios, difficulty variants, and individual SPs. Mean fidelity scores ranged from 4.40 to 4.58 across the four scenarios and from 4.49 to 4.55 across the two difficulty variants. Scores across the five individual SPs ranged from 4.20 to 4.85 (Figure 2), suggesting that no single scenario, difficulty variant, or SP drove the overall result. Scores grouped by MT showed a wider range, from 3.92 to 4.88, likely reflecting differences in rater strictness rather than SP portrayal quality. Each subgroup contained few encounters, so all four subgroup comparisons were interpreted descriptively. Internal consistency was 0.86 for the five portrayal items and 0.57 for the four interview-quality items.

**Figure 2.**
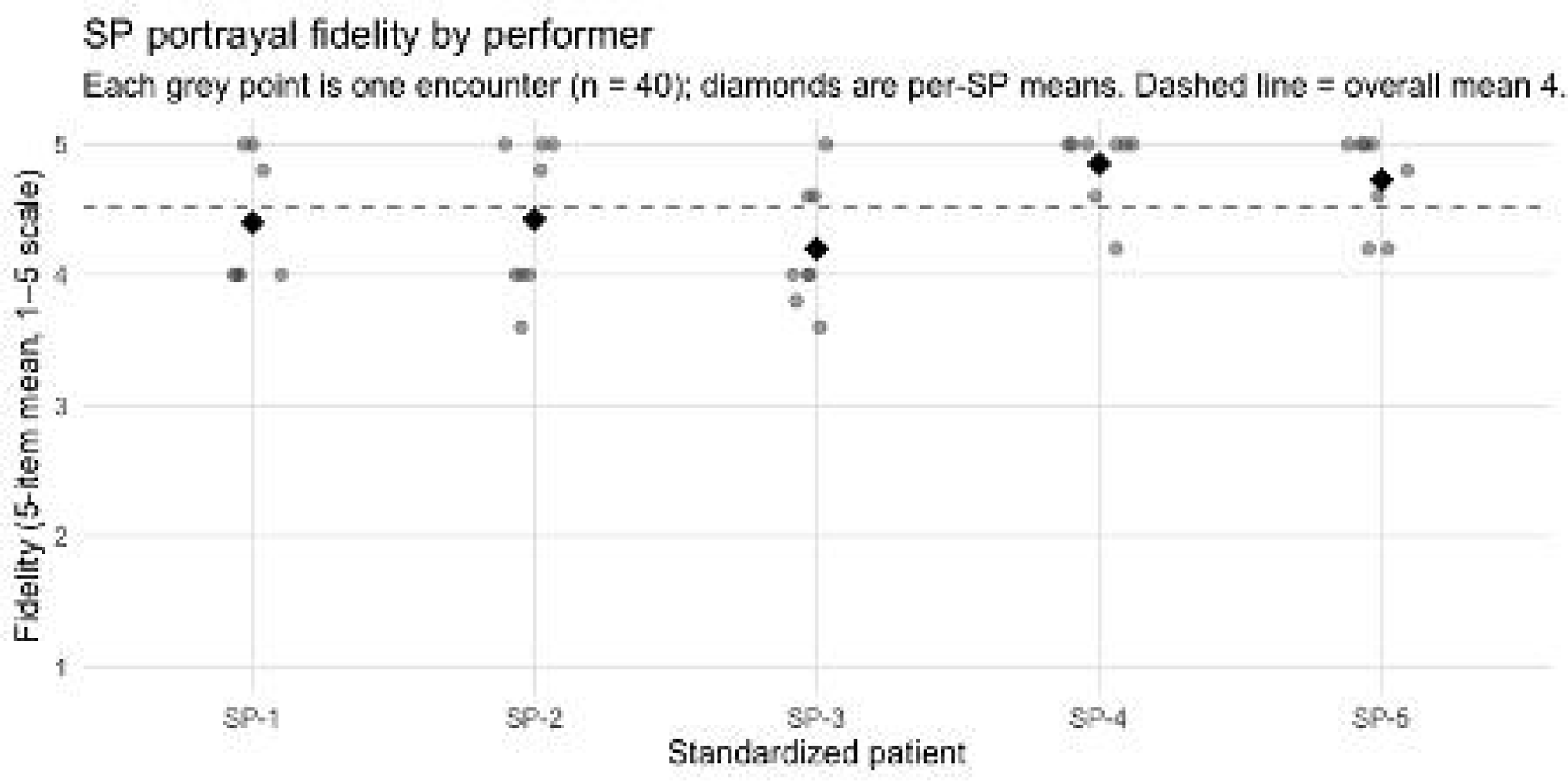
SP portrayal fidelity by performer. Each gray point represents one encounter (n = 40). Diamonds show the mean for each SP, and the dashed line shows the overall mean of 4.52. Fidelity was calculated as the mean of the five portrayal items using the 1-to-5 training scale

Note that the Phase 1 instruments used 1-to-5 scales, except the MT case-difficulty item, which had four options, whereas Phase 2 and Phase 3 used a 0-to-6 scale; ratings from Phase 1 were not pooled with ratings from the later phases.

SPs recorded only minor anomalies in the three free-text prompts. They noted two household interruptions from a voice assistant activating, one case in which the time limit prevented the MT from completing all questions, and several instances of the MT asking for the physician’s name, which an intake encounter would not require. One SP flagged a suicide-screening question on the women’s-health case as unexpected.

### Phase 2. Chatbot performance across case difficulty

All 80 conversations in the design yielded complete surveys and were analyzed. SPs and difficulty levels were equally represented, with 40 Easy and 40 Difficult conversations and 16 from each of the five SPs.

There were 1,000 element-level judgments for each of the two checklist outcomes: whether the chatbot asked about the element, and whether it gave the SP enough opportunity to address the element satisfactorily. The chatbot asked about the critical clinical element 59.3% of the time in Easy variants and 50.2% of the time in the Difficult variants. After accounting for differences between the four clinical cases, the odds that the chatbot asked about an element were 1.49 times higher in Easy variants than in Difficult variants (95% CI, 1.15 to 1.93; p = .0023). SPs reported satisfactory opportunity to address 73.3% of critical elements in Easy variants and 64.6% in Difficult variants. The adjusted odds of satisfactory coverage were 1.47 times higher in Easy variants (95% CI, 1.12 to 1.93; p = .0050).

The first sensitivity analysis fitted both outcomes as mixed-effects models with a random intercept for SP. The second removed the case adjustment. Both agreed with the primary models in direction and magnitude (Table 2).

**Table 2.** Element-level logistic regression models for Phase 2. Odds ratios compare Easy with Difficult variants. Values above 1 indicate that the outcome was more likely in Easy variants. Unadjusted models compare the two difficulty levels directly. Case-adjusted models account for differences across the four clinical cases and represent the primary analysis. The mixed-effects models allowed each SP to have a different baseline. Each element-level outcome includes 1,000 judgments. The logistic models used standard errors clustered by SP; the mixed-effects models instead included a random intercept for SP. The sentiment model used conversation-level clustering and was not adjusted for case.

| **Model** | **Odds ratio** | **95% CI** | **p** |
| --- | --- | --- | --- |
| Element asked, unadjusted | 1.45 | 1.17 to<br>1.79 | 0.0006 |
| Element asked, case-adjusted (primary) | 1.49 | 1.15 to<br>1.93 | 0.0023 |
| Element asked, mixed-effects sensitivity | 1.55 | 1.19 to<br>2.02 | 0.0013 |
| Element addressed satisfactorily, unadjusted | 1.50 | 1.13 to<br>1.99 | 0.0049 |
| Element addressed satisfactorily, case-adjusted (primary) | 1.47 | 1.12 to<br>1.93 | 0.0050 |
| Element addressed satisfactorily, mixed-effects sensitivity | 1.53 | 1.15 to<br>2.04 | 0.0036 |
| Negative sentiment in free-text reflections | 1.08 | 0.47 to<br>2.50 | 0.855 |

The difference between Easy and Difficult scenarios varied by case, ranging from +16.5 to −1.5 points for asking and from +13.3 to −1.5 points for addressing. These differences were not limited to cases in which the checklist became longer.

Summary quality was uniformly high and did not differ appreciably by difficulty. The ten PDQI-10 items averaged 5.85 in Easy variants and 5.77 in Difficult variants. No individual item differed significantly between variants. Easy variants scored higher on seven of the ten items, were tied on three, and were lower on none. Ratings concentrated at the top of the scale, where the maximum accounted for 90% of Easy and 83% of Difficult ratings. The comparison of negative sentiment in the SPs’ free-text reflections was inconclusive (odds ratio 1.08; 95% CI, 0.47 to 2.50; p = .855).

#### SP Debriefing Interview

Intercoder reliability, estimated with Gwet’s AC2^21^, was 0.78, in the substantial range^24^. The findings are reported in Table 3.

**Table 3.**
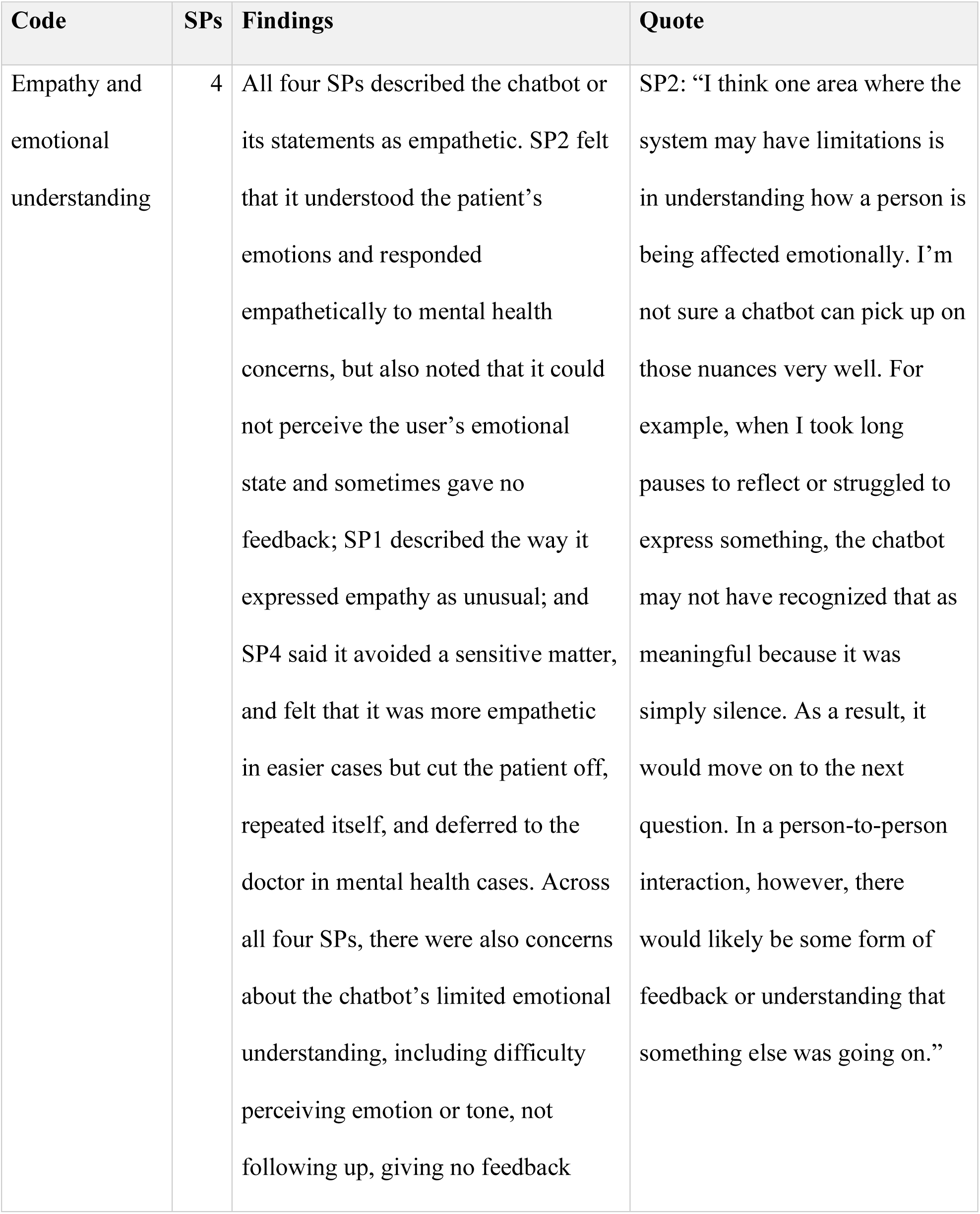
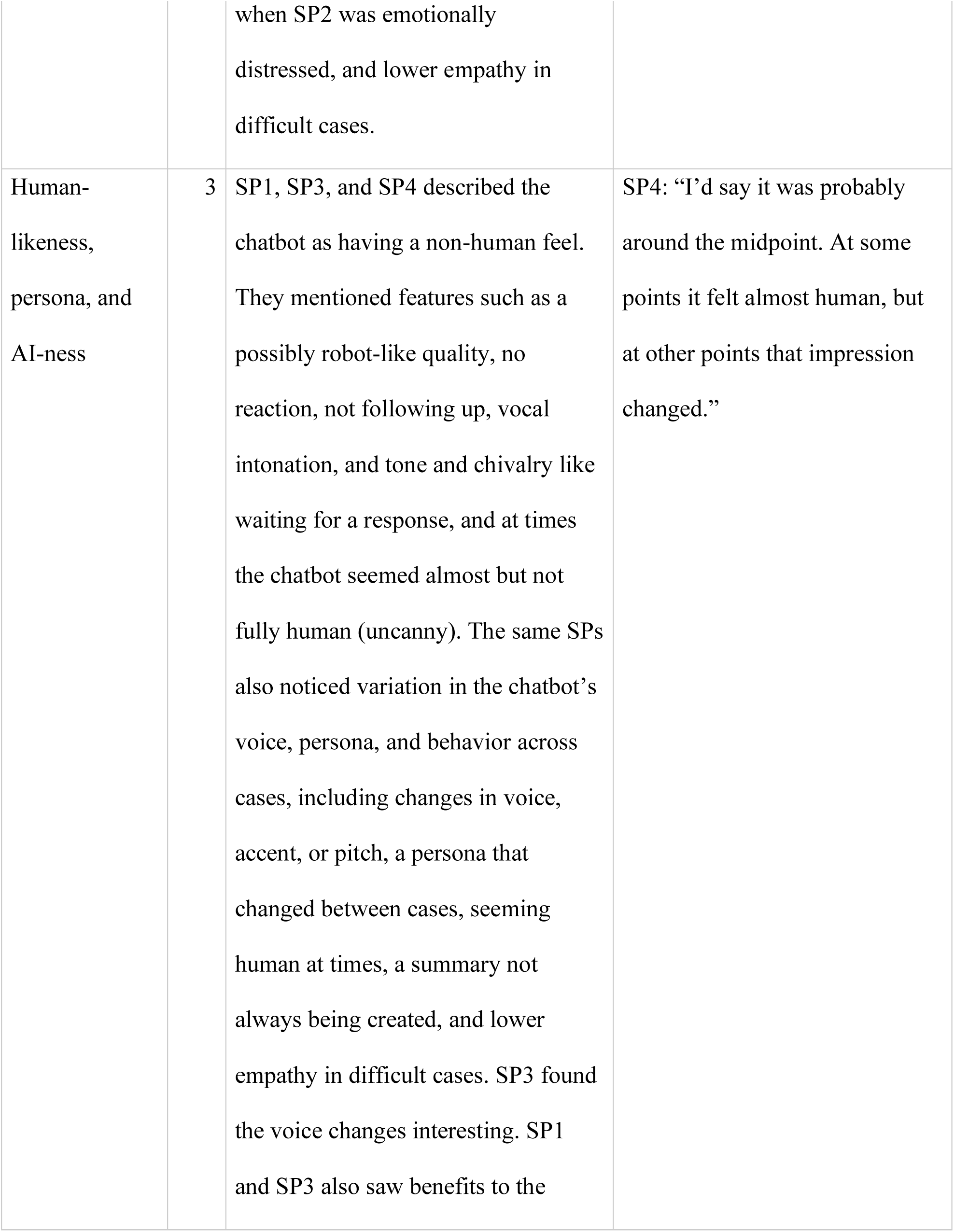
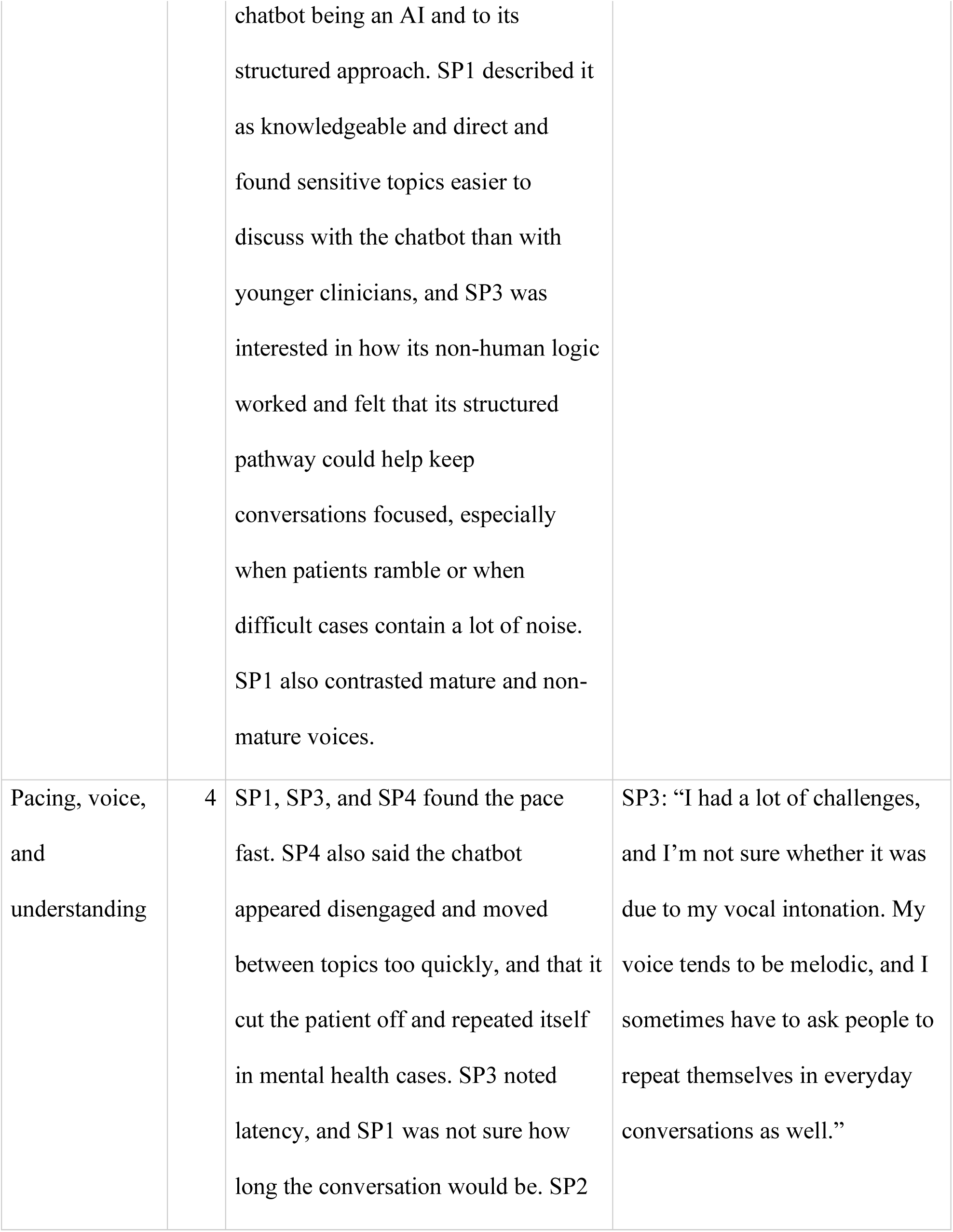
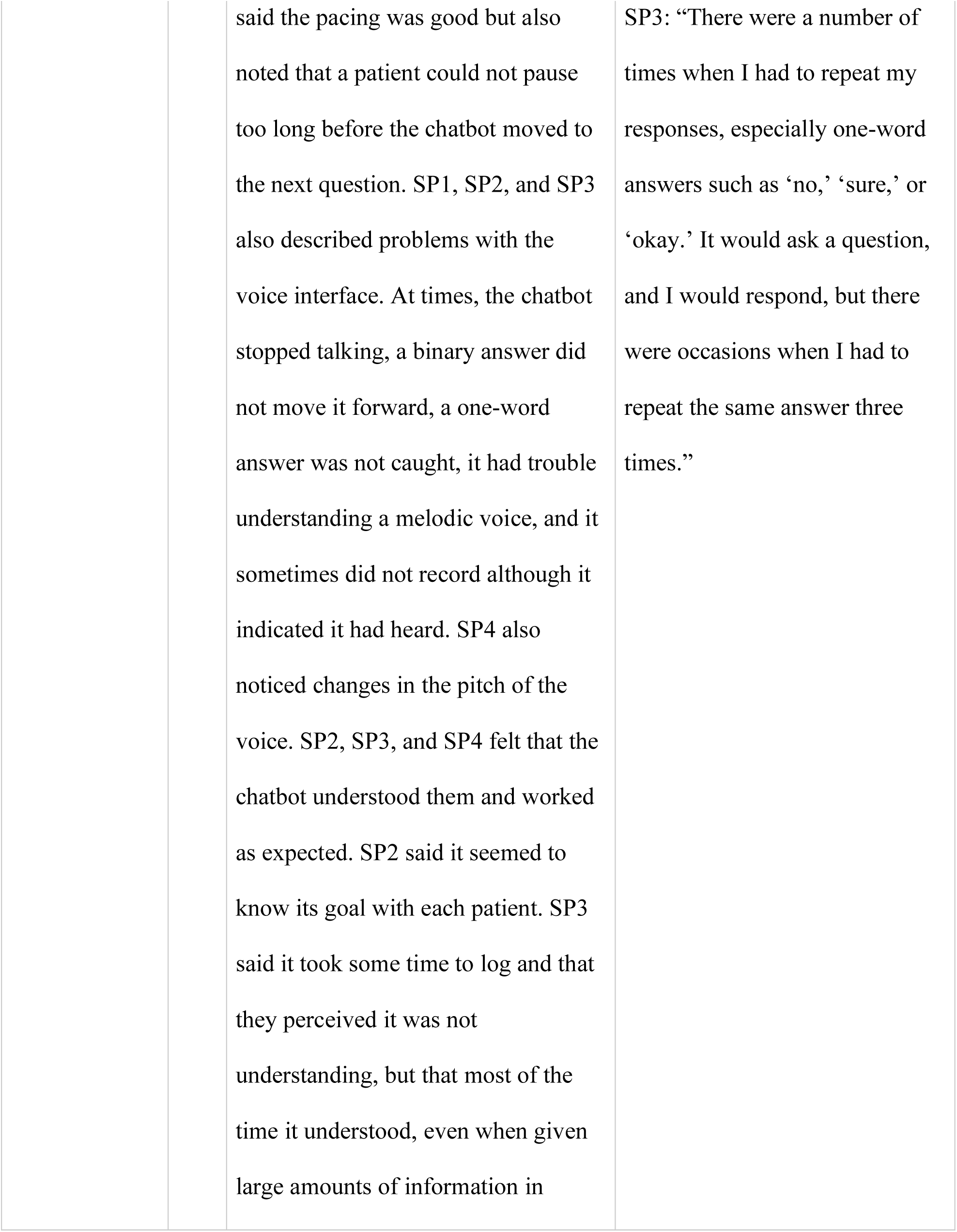
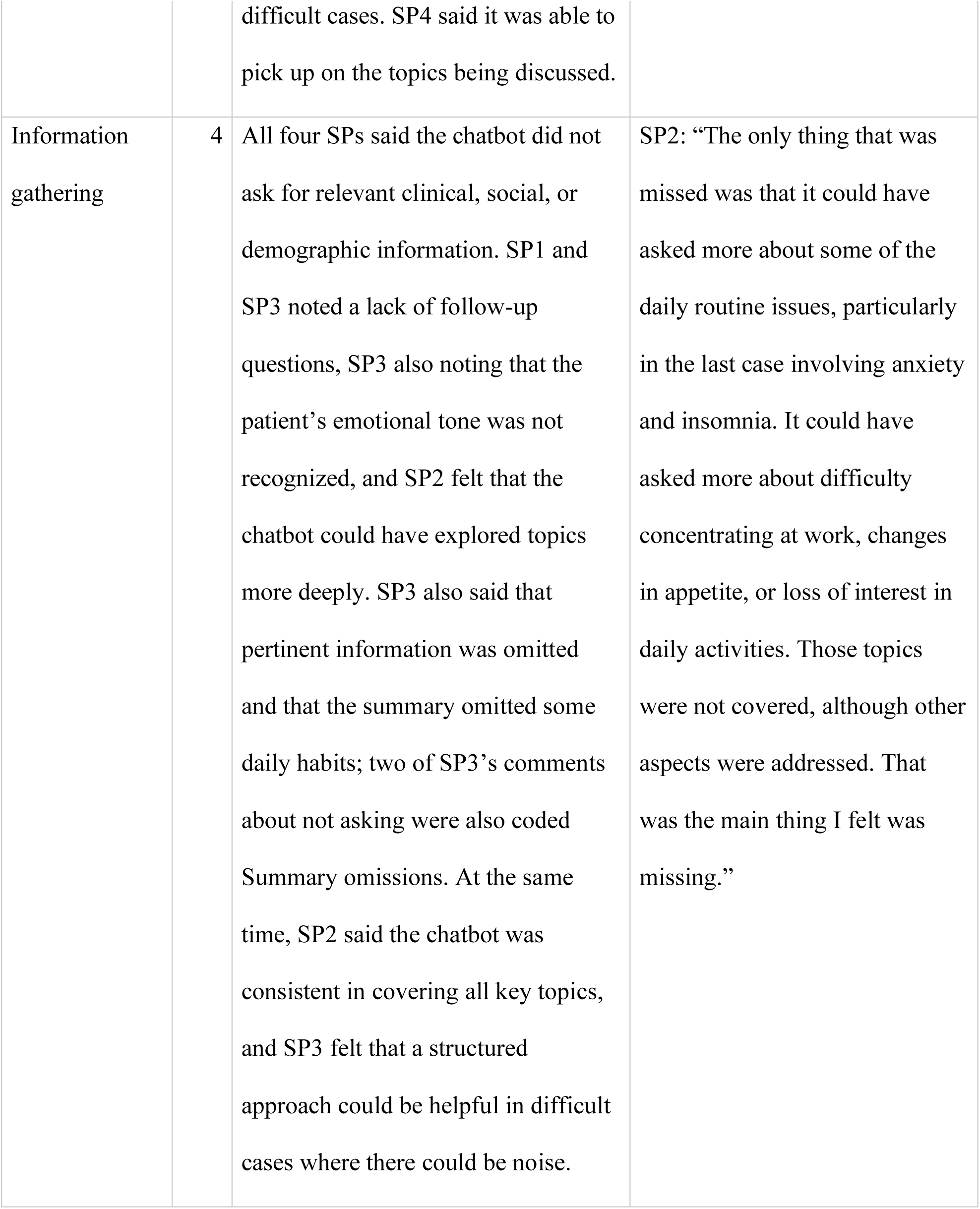
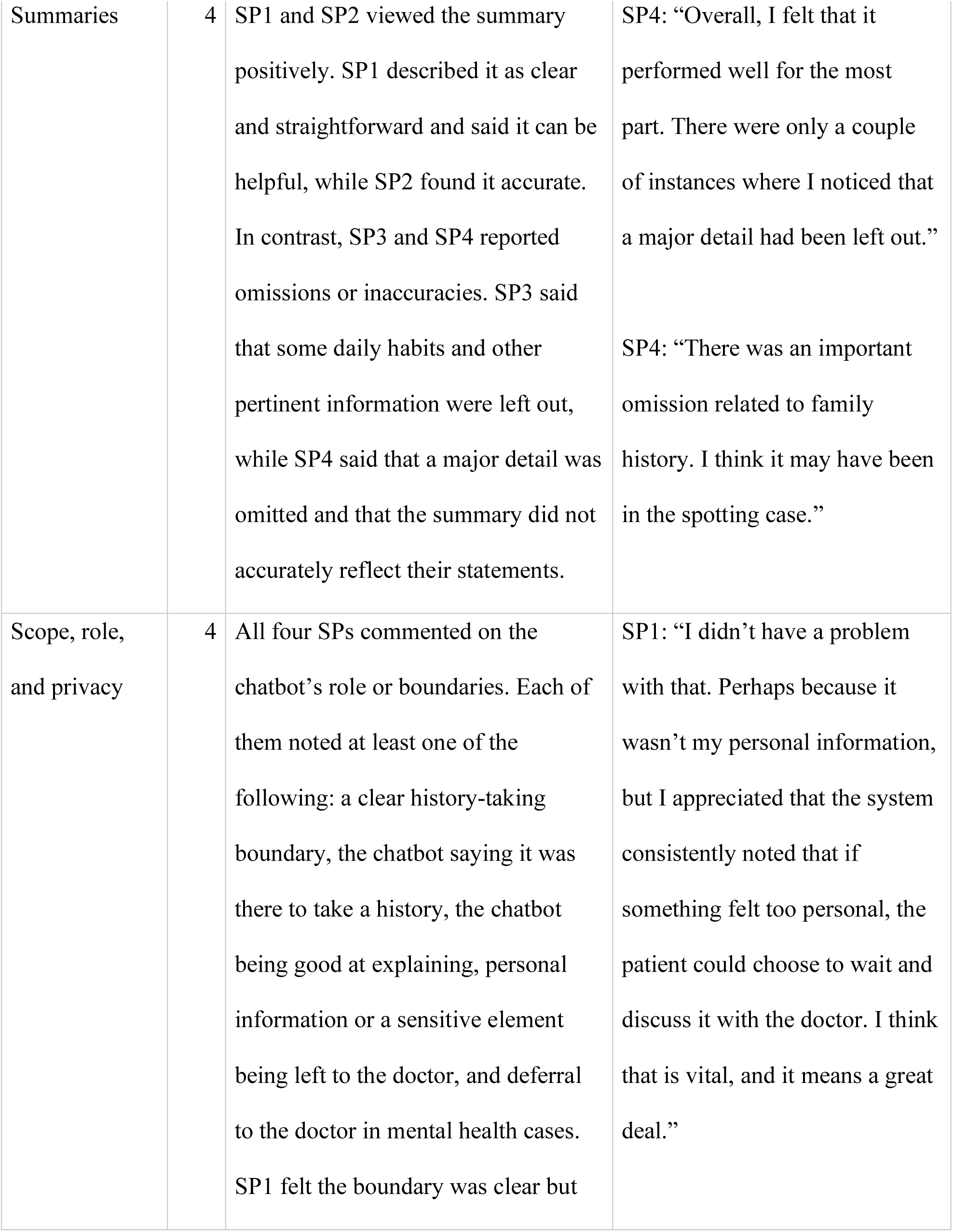
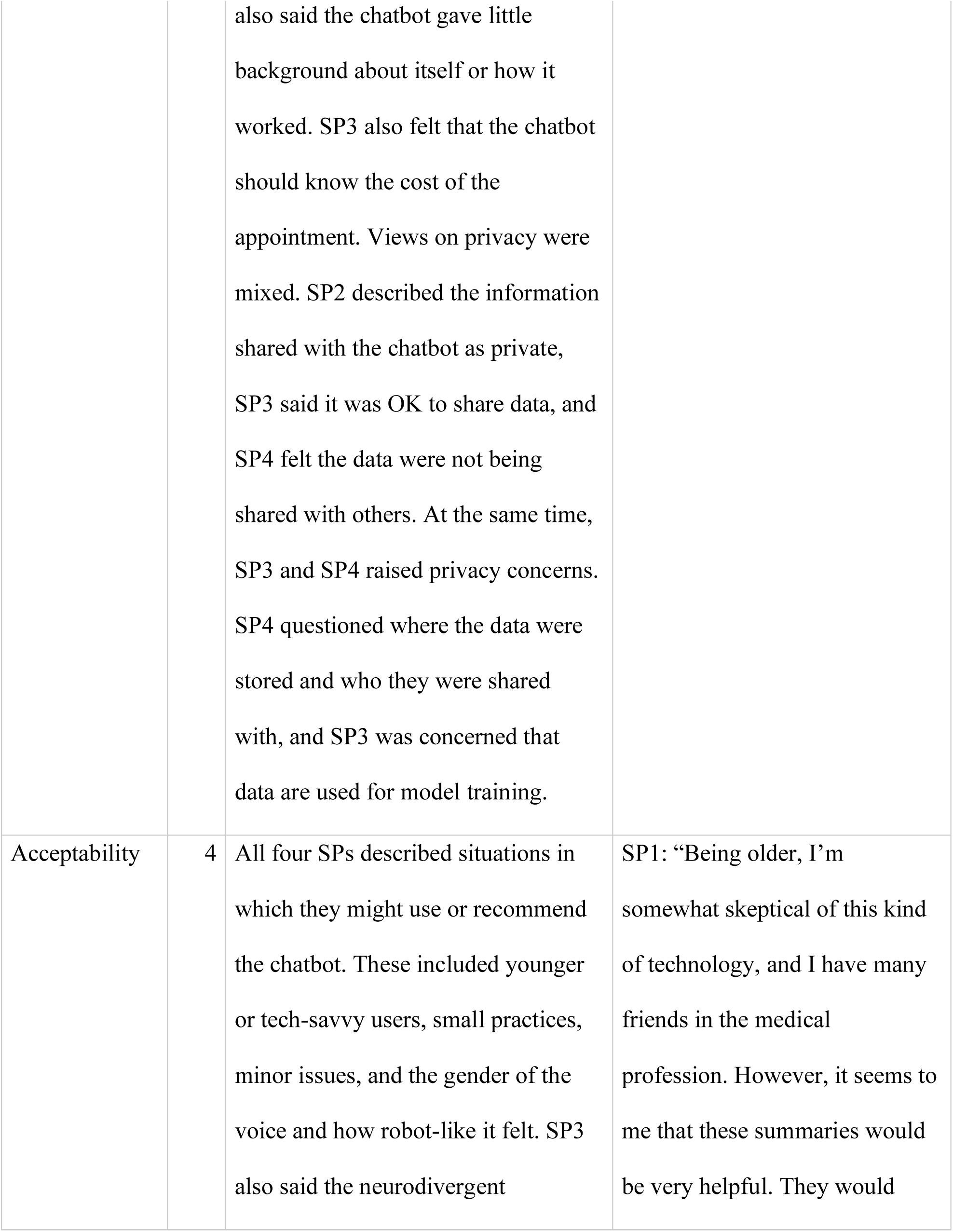
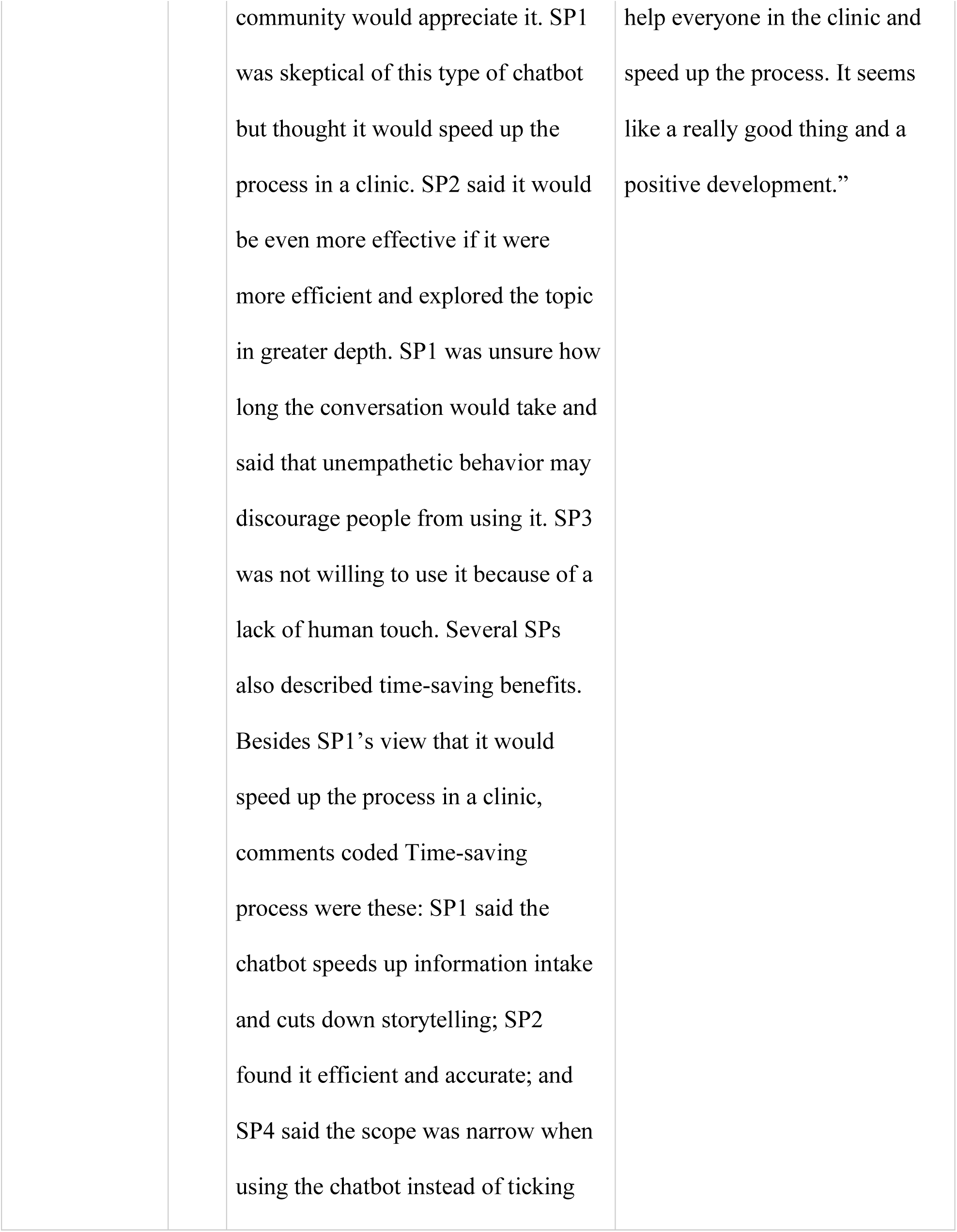
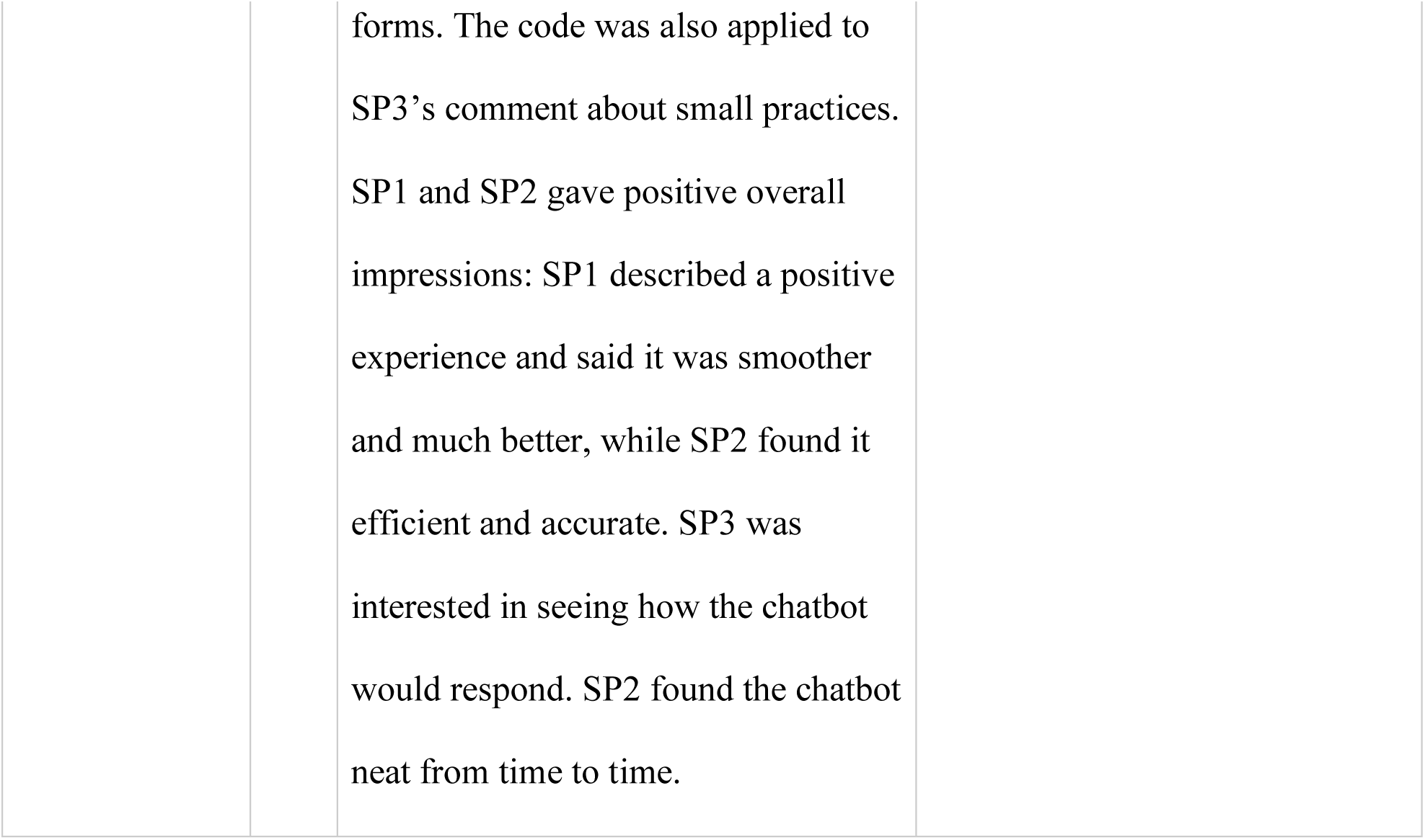
Interview Findings.

| Code | SPs | Findings | Quote |
| --- | --- | --- | --- |
| Empathy and emotional understanding | 4 | All four SPs described the chatbot or its statements as empathetic. SP2 felt that it understood the patient's emotions and responded empathetically to mental health concerns, but also noted that it could not perceive the user's emotional state and sometimes gave no feedback; SP1 described the way it expressed empathy as unusual; and SP4 said it avoided a sensitive matter, and felt that it was more empathetic in easier cases but cut the patient off, repeated itself, and deferred to the doctor in mental health cases. Across all four SPs, there were also concerns about the chatbot's limited emotional understanding, including difficulty perceiving emotion or tone, not following up, giving no feedback when SP2 was emotionally distressed, and lower empathy in difficult cases. | SP2: "I think one area where the system may have limitations is in understanding how a person is being affected emotionally. I'm not sure a chatbot can pick up on those nuances very well. For example, when I took long pauses to reflect or struggled to express something, the chatbot may not have recognized that as meaningful because it was simply silence. As a result, it would move on to the next question. In a person-to-person interaction, however, there would likely be some form of feedback or understanding that something else was going on." |
| Human-likeness, persona, and AI-ness | 3 | SP1, SP3, and SP4 described the chatbot as having a non-human feel. They mentioned features such as a possibly robot-like quality, no reaction, not following up, vocal intonation, and tone and chivalry like waiting for a response, and at times the chatbot seemed almost but not fully human (uncanny). The same SPs also noticed variation in the chatbot's voice, persona, and behavior across cases, including changes in voice, accent, or pitch, a persona that changed between cases, seeming human at times, a summary not always being created, and lower empathy in difficult cases. SP3 found the voice changes interesting. SP1 and SP3 also saw benefits to the<br>chatbot being an AI and to its structured approach. SP1 described it as knowledgeable and direct and found sensitive topics easier to discuss with the chatbot than with younger clinicians, and SP3 was interested in how its non-human logic worked and felt that its structured pathway could help keep conversations focused, especially when patients ramble or when difficult cases contain a lot of noise. SP1 also contrasted mature and non-mature voices. | SP4: "I'd say it was probably around the midpoint. At some points it felt almost human, but at other points that impression changed." |
| Pacing, voice, and understanding | 4 | SP1, SP3, and SP4 found the pace fast. SP4 also said the chatbot appeared disengaged and moved between topics too quickly, and that it cut the patient off and repeated itself in mental health cases. SP3 noted latency, and SP1 was not sure how long the conversation would be. SP2 | SP3: “I had a lot of challenges, and I’m not sure whether it was due to my vocal intonation. My voice tends to be melodic, and I sometimes have to ask people to repeat themselves in everyday conversations as well.” |
|  |  | said the pacing was good but also noted that a patient could not pause too long before the chatbot moved to the next question. SP1, SP2, and SP3 also described problems with the voice interface. At times, the chatbot stopped talking, a binary answer did not move it forward, a one-word answer was not caught, it had trouble understanding a melodic voice, and it sometimes did not record although it indicated it had heard. SP4 also noticed changes in the pitch of the voice. SP2, SP3, and SP4 felt that the chatbot understood them and worked as expected. SP2 said it seemed to know its goal with each patient. SP3 said it took some time to log and that they perceived it was not understanding, but that most of the time it understood, even when given large amounts of information in<br>difficult cases. SP4 said it was able to pick up on the topics being discussed. | SP3: “There were a number of times when I had to repeat my responses, especially one-word answers such as ‘no,’ ‘sure,’ or ‘okay.’ It would ask a question, and I would respond, but there were occasions when I had to repeat the same answer three times.” |
| Information gathering | 4 | All four SPs said the chatbot did not ask for relevant clinical, social, or demographic information. SP1 and SP3 noted a lack of follow-up questions, SP3 also noting that the patient's emotional tone was not recognized, and SP2 felt that the chatbot could have explored topics more deeply. SP3 also said that pertinent information was omitted and that the summary omitted some daily habits; two of SP3's comments about not asking were also coded Summary omissions. At the same time, SP2 said the chatbot was consistent in covering all key topics, and SP3 felt that a structured approach could be helpful in difficult cases where there could be noise. | SP2: "The only thing that was missed was that it could have asked more about some of the daily routine issues, particularly in the last case involving anxiety and insomnia. It could have asked more about difficulty concentrating at work, changes in appetite, or loss of interest in daily activities. Those topics were not covered, although other aspects were addressed. That was the main thing I felt was missing." |
| Summaries | 4 | SP1 and SP2 viewed the summary positively. SP1 described it as clear and straightforward and said it can be helpful, while SP2 found it accurate. In contrast, SP3 and SP4 reported omissions or inaccuracies. SP3 said that some daily habits and other pertinent information were left out, while SP4 said that a major detail was omitted and that the summary did not accurately reflect their statements. | SP4: “Overall, I felt that it performed well for the most part. There were only a couple of instances where I noticed that a major detail had been left out.”<br>SP4: “There was an important omission related to family history. I think it may have been in the spotting case.” |
| Scope, role, and privacy | 4 | All four SPs commented on the chatbot’s role or boundaries. Each of them noted at least one of the following: a clear history-taking boundary, the chatbot saying it was there to take a history, the chatbot being good at explaining, personal information or a sensitive element being left to the doctor, and deferral to the doctor in mental health cases. SP1 felt the boundary was clear but<br>also said the chatbot gave little background about itself or how it worked. SP3 also felt that the chatbot should know the cost of the appointment. Views on privacy were mixed. SP2 described the information shared with the chatbot as private, SP3 said it was OK to share data, and SP4 felt the data were not being shared with others. At the same time, SP3 and SP4 raised privacy concerns. SP4 questioned where the data were stored and who they were shared with, and SP3 was concerned that data are used for model training. | SP1: “I didn’t have a problem with that. Perhaps because it wasn’t my personal information, but I appreciated that the system consistently noted that if something felt too personal, the patient could choose to wait and discuss it with the doctor. I think that is vital, and it means a great deal.” |
| Acceptability | 4 | All four SPs described situations in which they might use or recommend the chatbot. These included younger or tech-savvy users, small practices, minor issues, and the gender of the voice and how robot-like it felt. SP3 also said the neurodivergent | SP1: “Being older, I’m somewhat skeptical of this kind of technology, and I have many friends in the medical profession. However, it seems to me that these summaries would be very helpful. They would |
|  |  | community would appreciate it. SP1 was skeptical of this type of chatbot but thought it would speed up the process in a clinic. SP2 said it would be even more effective if it were more efficient and explored the topic in greater depth. SP1 was unsure how long the conversation would take and said that unempathetic behavior may discourage people from using it. SP3 was not willing to use it because of a lack of human touch. Several SPs also described time-saving benefits. Besides SP1's view that it would speed up the process in a clinic, comments coded Time-saving process were these: SP1 said the chatbot speeds up information intake and cuts down storytelling; SP2 found it efficient and accurate; and SP4 said the scope was narrow when using the chatbot instead of ticking<br>forms. The code was also applied to SP3's comment about small practices. SP1 and SP2 gave positive overall impressions: SP1 described a positive experience and said it was smoother and much better, while SP2 found it efficient and accurate. SP3 was interested in seeing how the chatbot would respond. SP2 found the chatbot neat from time to time. | help everyone in the clinic and speed up the process. It seems like a really good thing and a positive development.” |

### Phase 3. Reviewer agreement

Of the 120 encounters, 19 were not assigned for review because one or both documents were missing, leaving 101 encounters, 32 resident-led, 69 chatbot-led. Eighteen of the 19 losses were technical capture failures in identifiable sessions, and the eight unusable resident-led transcripts all involved the same SP. One reviewer was assigned 61 encounters and the other 60. Of the encounters assigned to both reviewers, 15 were chatbot-led, and 5 were resident-led. Each of the four cases accounted for roughly one-quarter of encounters in both groups. The resident-led group included exactly 8 encounters per case, while the chatbot-led group included 17, 19, 17, and 16 encounters across the four cases, respectively.

The reliability sample provided 260 critical-element judgments. Reviewers showed moderate agreement on whether an element was raised (AC1 = 0.49; 95% CI, 0.39 to 0.60; 74.6% agreement) based on the Landis and Koch benchmarks^24^. Agreement on whether an element was addressed adequately was substantial across all 260 judgments (AC1 = 0.68; 95% CI, 0.55 to 0.79; 83.8% agreement). Among the 171 judgments where at least one reviewer marked the element as asked, agreement was similar (AC1 = 0.70; 95% CI, 0.52 to 0.84; 80.1% agreement). Of the 260 judgments, 89 concerned elements that neither reviewer considered raised; for these, reviewers agreed 91% of the time that the element was not addressed adequately (AC1 = 0.90; 95% CI, 0.83 to 0.96). Because agreement on elements that were never raised is less informative, we treated the asked-only estimate as the more interpretable measure.

Absolute agreement on the two ordinal totals was low, with ICC(A,1) of 0.10 (95% CI, −0.09 to 0.38) for the summary total and 0.21 (95% CI, −0.12 to 0.55) for the transcript total. Reviewer 1 scored higher on 12 of the 15 items, by a mean of 0.86 points on summary items and 0.44 on transcript items (Figure 3); the primary model put the gap at 7.9 points on the summary total and 1.8 on the transcript total (both p < .001). Reviewers gave identical scores on 23.5% of summary and 21.0% of transcript ratings and were within one point on 67.0% and 71.0%, respectively. Item-level AC2 averaged 0.84 for summary items and 0.87 for transcript items (range 0.68 to 0.98); for Risk of patient harm it was 0.80 (95% CI, 0.63 to 0.97), with identical scores on 30% of ratings.

**Figure 3.**
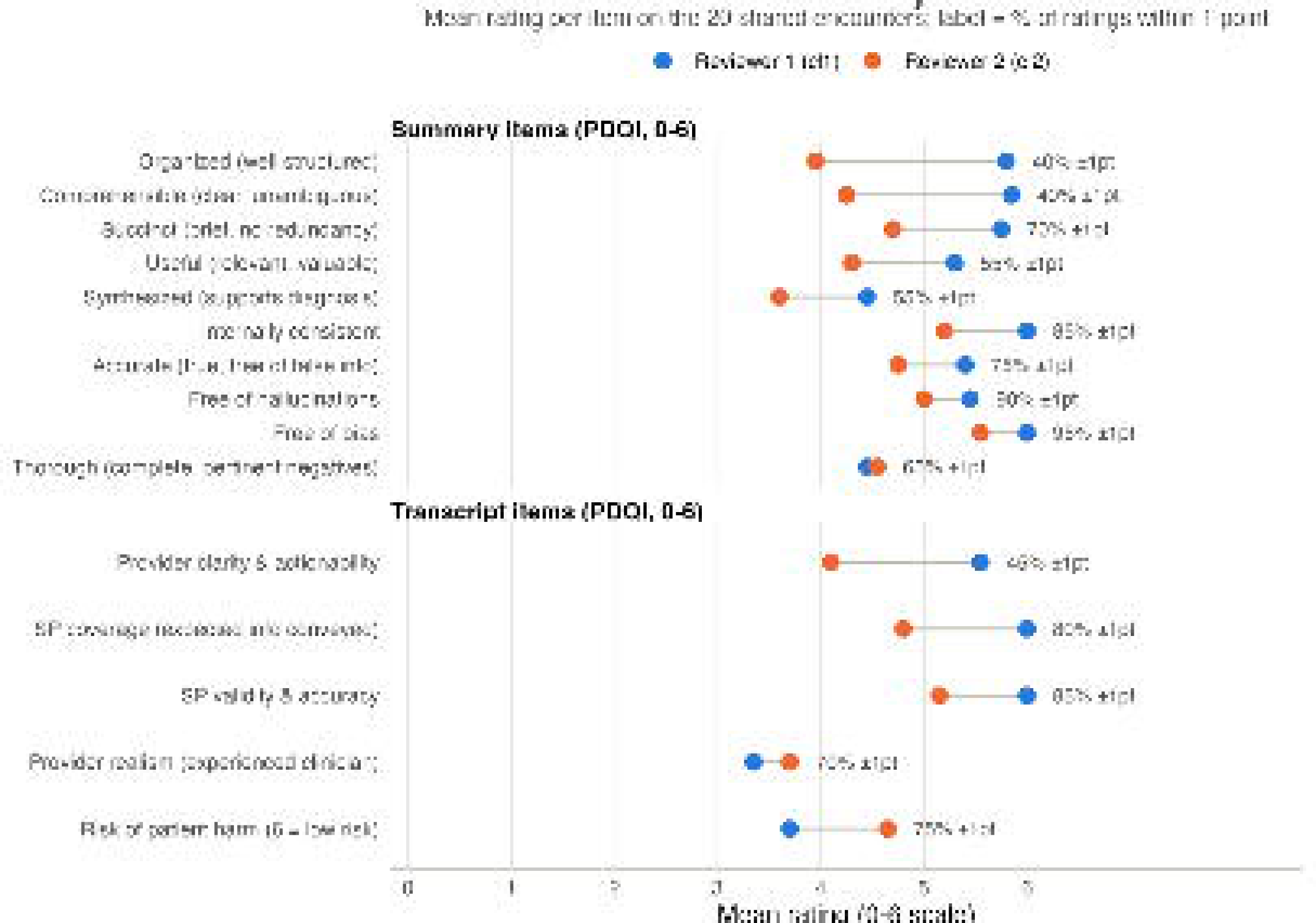
Reviewer calibration by item. Points show the mean rating for each item across the 20 encounters rated by both reviewers, shown separately for the ten summary-quality items and the five transcript-quality items. The percentage shown for each row indicates how often the two reviewers’ ratings were within one point of each other

### Phase 3. Resident-led versus chatbot-led encounters

Adjusted differences compare resident-led with chatbot-led encounters, after accounting for reviewer and case variant. A positive value means the resident-led group scored higher. Neither the written summary nor the encounter itself differed in rated quality between the two groups (Table 4). Residents asked a significantly larger proportion of each case’s critical elements and addressed a larger share of these satisfactorily than did the chatbot.

**Table 4.** Comparison of resident-led and chatbot-led encounters across four outcomes (101 encounters; 121 reviewer-by-encounter records). Group values are shown as encounter-level mean ± SD. Adjusted differences are calculated as resident-led minus chatbot-led and account for reviewer and case variant. Positive values indicate higher scores for resident-led encounters. For the summary total, the unadjusted means are higher for chatbot-led encounters but the adjusted difference is positive, because the two reviewers differed in leniency and rated the two groups in different proportions.

| Outcome | Resident | Chatbot | Adjusted difference | 95% CI | p |
| --- | --- | --- | --- | --- | --- |
| PDQI-10 summary total (0–60) | 49.92 ± 4.02 | 50.75 ± 5.54 | +0.415 | –1.527 to 2.356 | 0.670 |
| Transcript total (0–30) | 24.88 ± 2.86 | 23.99 ± 2.85 | +1.020 | –0.102 to 2.141 | 0.074 |
| Proportion of elements asked | 0.67 ± 0.18 | 0.52 ± 0.18 | +0.125 | 0.070 to 0.181 | <0.001 |
| Proportion addressed satisfactorily | 0.64 ± 0.14 | 0.54 ± 0.14 | +0.092 | 0.038 to 0.146 | 0.001 |

The descriptive comparison averaged the two reviewers within each encounter and reached the same conclusion as the primary model on all four outcomes. Hedges’ g was 0.86 for elements asked and 0.72 for elements addressed satisfactorily, against −0.16 for the summary total and 0.31 for the transcript total. Case-adjusted contrasts that omit the reviewer term gave −0.78 (95% CI, −2.98 to 1.42; p = .4844) for the summary total, +0.94 (95% CI, −0.21 to 2.09; p = .1087) for the transcript total, +0.15 (95% CI, 0.08 to 0.22; p < .001) for elements asked, and +0.10 (95% CI, 0.05 to 0.16; p < .001) for elements addressed satisfactorily.

In the clinician reviewers’ ratings, Difficult variants showed lower coverage than Easy variants, at 0.52 versus 0.61 for elements asked and 0.55 versus 0.59 for elements addressed satisfactorily, while the summary total and transcript total were nearly identical.

The joint test across all 15 items showed that the two groups differed somewhere within the item set (*χ²(*15) = 77.6, *p* < .001). The magnitude of this difference also varied significantly across items (*χ²*(14) = 76.7, *p* < .001). A single summary contrast across items would therefore not adequately represent the data. The average item difference was +0.10 (95% CI, −0.05 to 0.24; p = .20).

The Risk of patient harm item was the only item to differ significantly after Bonferroni correction. Resident-led encounters scored higher on this item, which indicates lower perceived risk (+0.97, 95% CI, 0.49 to 1.46; corrected p = .003). The remaining 14 differences fell between −0.30 and +0.57 points and none reached significance after correction. All corrected p values for those items were .19 or greater. The largest were provider realism at +0.57 and SP coverage at −0.30. Within the transcript scale, the three provider-rated item differences summed to +1.59 points and the two SP-rated item differences to −0.43 points. Neither adjustment could be treated as common across items: the reviewer effect (χ²(14) = 319.8, p < .001) and the difficulty effect (χ²(14) = 25.4, p = .03) both varied by item. Refitting with item-specific reviewer terms, and again with item-specific reviewer and difficulty terms, left Risk of patient harm as the only item significant after correction, at +0.81 and +0.80.

## Discussion

### Principal Findings

We developed and tested an SP-based method to evaluate how well a patient-facing AI chatbot collects an initial clinical history. We conducted the study in three linked phases. We demonstrated the feasibility of using SPs to evaluate AI chatbots. Importantly, the findings suggest that conversational intake tools should be evaluated separately in three areas: the information captured during the conversation, measured with a critical-element checklist; how the encounter was conducted and experienced by the patient, captured in the SP debriefing interviews; and the quality of the resulting documentation, rated with the PDQI-10.

The clearest contrast in our results was between coverage and quality. The summary and transcript totals were high in every group. The summary total did not differ between groups, and the transcript total did not reach significance. In contrast, coverage of the critical clinical elements clearly separated human from AI, with a large effect for elements asked (g = 0.86) and a moderate-to-large effect for elements addressed (g = 0.72).

Critical clinical elements that were never discussed will not appear in the summary. A measure applied only to the summary therefore evaluates the quality of the documentation rather than the completeness of the conversation that produced it. Such omissions can be identified only from the conversation itself, through the transcript or the patient’s own report.

This study assessed coverage using two independent sources. SPs completed a structured assessment after each conversation. The blinded clinician reviewers also assessed the transcripts. Both measures showed lower coverage in the Difficult variants, although in the SP checklists, the gap varied across the four cases and reversed slightly in one case for each outcome. This consistency suggests that the observed difference did not depend on a single rating source. The difficulty labels worked as intended, which supported adjusting the Phase 3 comparison for case variant. Difficulty was created in two ways. In two scenarios, the checklist stayed the same, so the difference came only from how the case was portrayed. In the other two, the Difficult versions included eight additional critical elements in total (one in Case 3 and seven in Case 4). Coverage was lower in Difficult cases both when the checklist stayed the same and when it became longer. This suggests that the lower coverage was not simply caused by having more checklist items.

The SP debriefing interviews revealed a similar pattern from the patient perspective. All four SPs reported that the chatbot missed relevant clinical, social, or demographic information, and two noted that the final summary omitted or misstated information they had shared. They also described conversational behaviors that may have contributed to these gaps, including moving too quickly, shifting topics before completing the previous one, interrupting the patient, and failing to follow up on brief responses. Views on empathy were mixed, although some SPs felt the chatbot was less empathetic in the more difficult cases. These findings reflect the SPs’ direct experience of the encounter and suggest that conversational conduct should be evaluated separately from information capture and documentation quality.

When the 15 rating items, ten summary-quality and five transcript-quality, were compared individually, a difference emerged that the transcript total did not reflect. Reviewers rated resident-led encounters as having less potential for patient harm. This was the only one of the 15 items that remained significant after correction and was one of the five items included in the transcript total. The two reviewers showed substantial agreement on this item, even though they gave exactly the same score on fewer than one-third of the ratings. This finding therefore requires replication before supporting conclusions about relative safety. The transcript scale combines two different types of items. Three items rate the provider, either the chatbot or the resident, on realism, clarity, and risk of patient harm. The other two items rate how well the SP portrayed the case. These two sets of items moved in opposite directions. In the item-level model, the provider items favored the resident group by 1.59 points, while the SP items favored the chatbot group by 0.43 points. As a result, the overall null difference does not mean the two groups performed the same. Instead, the difference in provider performance was partly offset by the SP portrayal items.

Rater strictness varied. Mean scores differed by nearly one full point between the strictest and most lenient MTs in Phase 1. The two clinician reviewers in Phase 3 differed in the same direction on most items, and that per-item gap accumulated across the scale totals. However, the reviewers were within one point of each other on about seven of every ten ratings. This pattern suggests that much of the difference was one of rating leniency. Absolute-agreement statistics alone may therefore give an incomplete picture, while coefficients that give near-full credit to one-point differences may give an overly favorable one. Evaluations of this kind should report calibration alongside agreement. Analyses should also account for rater identity when raters are unevenly distributed across comparison groups.

### Comparison With Prior Work

These results add to prior work in several ways. The chatbot in this study asked about 50% to 60% of the critical clinical elements, and SPs reported satisfactory opportunity to address 65% to 73% of all critical elements. In a prior study in which physicians and medical students played the patients through a web-based conversational interface, the agentic system captured relevant history elements with accuracy and F1 scores above 85%, judged against gold-standard case histories.^5^ However, the chatbots, the people playing the patients, the delivery channels, the rubrics, and the metrics all differed between the two studies. In our study, trained SPs spoke with the chatbot by voice, and capture was measured as the proportions of critical elements asked and addressed satisfactorily. Any measure of information capture should therefore be reported alongside the rubric, the metric, and the delivery channel that produced it.

A recent simulation study evaluated a history-taking chatbot against case-specific checklists using six simulated patients and 66 primary care cases.^26^ Content capture ranged from 51% to 72% across chatbot configurations and varied by symptom type. That design varied the chatbot and held the cases constant, whereas our study held the chatbot constant and varied case difficulty. Importantly, we examined how closely SPs followed the written scripts, compared chatbot-led with resident-led encounters, and independently rated the resulting clinical documentation. A pilot study in a vestibular clinic reported 88.7% completion of prespecified history items across 19 patients, with migraine items under-ascertained.^27^ That system followed a 14-item structured questionnaire within a single symptom domain, whereas the chatbot in this study held open conversations, guided by the written rules used to train it, across four complaint types. Checklist coverage may therefore depend on how the history-taking is structured, the range of clinical topics, and the difficulty of the case.

Related work rates history-taking as one dimension of consultation quality, using scales adapted from established examination rubrics.^14^ A simulated-patient study in emergency medicine rated ChatGPT above physicians on history-taking, using a five-point scale applied to text-based consultations.^28^ Our study instead recorded whether each critical clinical element was asked; this measure favored the resident-led encounters. A rated impression of history-taking and a count of elements elicited measure different things.

### Limitations

This study has limitations. First, the two groups were formed by study phase rather than random assignment. The resident-led group contributed only 32 of the 101 encounters. This comparison should therefore be considered exploratory rather than a test of relative performance. The residents were physicians in training who had reviewed the written rules used to train the chatbot, so although they acted as intake clinicians, they do not represent the intake nurses whose role the chatbot plays. Second, the SPs are trained team members rather than actual patients, and SP portrayal is an accepted stand-in for patient presentation rather than the same thing^11,17^. The SPs knew which difficulty variant they were portraying when they rated coverage, although the reviewers, who were not told the difficulty variant, also found lower coverage in the Difficult variants. Third, 19 of the 120 encounters never reached review, and the eight unusable resident-led transcripts all involved a single SP, so an effect specific to that performer cannot be separated from the group difference. Fourth, the two reviewers agreed poorly on the ordinal totals in absolute terms, and the documentation ratings clustered in the upper part of their range, which together limit confidence in the two quality comparisons even after adjustment for reviewer. Agreement was stronger for the binary coverage judgments than for the ordinal total scores, so the coverage estimates are more strongly supported. For the item-level comparison, the initial model assumed that the effects of reviewer and case difficulty were the same across all 15 items. Both assumptions were rejected, although the refits reported above left the item-level conclusion unchanged. The 20 double-rated encounters were unevenly distributed across the eight case variants, with no ratings for one variant and no more than four for any other variant. Reliability therefore could not be estimated separately by case variant. Fifth, internal consistency was modest for the four interview-quality items, which were therefore used descriptively rather than as a scale. The analysis of negative sentiment was underpowered, with an interval spanning a halving to a 2.5-fold increase in the odds, so the comparison of SP experience across difficulty remains unresolved. The debriefing findings came from four SPs, each interviewed once, and are therefore exploratory. The case set was also narrow. Four chief complaints were used, the difficulty contrast was modest, and the SPs delivered the scripts clearly, although one SP reported that the chatbot had trouble understanding their melodic voice. Cases built to stress communication, through accent, highly emotional content, or limited English proficiency, and cases with genuinely ambiguous presentations would test the method under conditions this study did not include. The SP encounters relied throughout on videoconferencing and electronic rating forms, and on the chatbot in Phase 2, and the anomalies the SPs recorded in Phase 1 included two interruptions from a household voice assistant. A brief technical rehearsal and a simple interface should therefore be included in the protocol from the outset. Finally, the method was applied to a single chatbot, so the performance findings describe that tool rather than patient-facing conversational AI in general. However, the purpose of this study was to develop and initially validate the methods of SP-based chatbot evaluation and, for this, the findings are quite promising.

### Future Directions

The immediate next step is the full study that this phase was designed to support. The method will be applied to compare chatbot documentation with clinician documentation and to measure how patients experience the interaction. Extension to actual patients and clinicians would follow, consistent with recent real-world chatbot trials.^4^ Future work should address two questions. First, does coverage of the required clinical content affect later clinical decisions or actions? Second, do patients’ own ratings of the encounter provide information that reviewer-based scoring does not capture? Standardized methods for evaluating patient-facing conversational AI before clinical use remain limited.^29^ A remote method that requires only a small number of trained team members could make this type of evaluation practical. Patient-facing AI systems are already being used to collect clinical histories. Methods that identify what these conversations miss should therefore be used before deployment rather than after.

## Conclusions

Using the SP-based method, SPs delivered the written scripts of primary care visits with high fidelity across four chief complaints and two difficulty levels. The method also detected lower chatbot coverage as case difficulty increased. Coverage of the critical clinical elements clearly separated chatbot-led from resident-led encounters. In contrast, neither the summary total nor the transcript total separated the groups, although the transcript total numerically favored the resident-led encounters. Evaluations of patient-facing intake tools will need to measure what the conversation captures directly. Ratings of the resulting documentation alone may not identify important information that was missed during the encounter.

## Data Availability

All data produced in the present study are available upon reasonable request to the authors

## Acknowledgements

The authors wish to acknowledge the important contributions of our Standardized Patients – Deanna Haynes, Michael Flynn, Marci Hall, Keenan Payne, Alice Raver, and Darci Wantiez – and our Medical Testers – Chiara Bettale, MD, Lauren Chan, MD, Hudson Holmes, MD, Dheera Reddy, MD, Bry Reinhardt, MD, MPH, and Tyler Schuhert, MD. We also acknowledge Dr. Trent Rosenbloom for reviewing the original case vignettes.

